# Myocardial Tug-of-War Is a Determinant of Left Ventricular Function and Failure

**DOI:** 10.64898/2026.04.23.26351629

**Authors:** Markus B. Harbo, Mohammad Javad Sadeghinia, Yasmin Donnabel M. Reyes, Radostin D. Simitev, Jia Li, Kjersti B. Blom, Tryggve H. Storås, Vigdis Rosseland, Nils Einar Kløw, Mathis K. Stokke, Kaspar Broch, Samuel T. Wall, Joakim Sundnes, Jon Arne Birkeland, Geir Øystein Andersen, William E. Louch, Godfrey L. Smith, Ivar Sjaastad, Emil K. S. Espe

## Abstract

**Background:** Heart failure with reduced ejection fraction is a leading cause of death worldwide, characterized by impaired left ventricular systolic function. Contractile, structural, and electrophysiological changes underpin this impairment, but how these changes collectively determine ventricular function remains unclear. We hypothesize that their integrated action involves a complex mechanical interplay at the myocardial mesoscale level, intermediate between individual cardiomyocytes and the global left ventricle.

**Methods:** We acquired high-resolution magnetic resonance images of healthy individuals and patients with myocardial infarction, and developed an analytical method to characterize *in vivo* contraction patterns in millimeter-sized myocardial units (i.e., at the mesoscale). Furthermore, we employed computational models to examine how mesoscale contraction patterns relate to the contraction mechanism, structure, and electrophysiology of the left ventricle.

**Results:** At the left ventricular mesoscale, we observed that weakly contracting myocardial units are transiently elongated by the contraction of adjacent, more strongly contracting units. These mesoscale interactions generate a “tug-of-war” that pervades the left ventricle in healthy hearts and becomes particularly prominent following myocardial infarction. This behavior is macroscopically invisible as the contraction patterns of opposing units cancel each other out, but it nevertheless shapes the efficiency of mechanical performance. In the healthy heart, recruitment of more uniformly contracting units (i.e., reduction in tug-of-war) supports augmented contractility during acute stress. However, following myocardial infarction, excessive tug-of-war contributes to impaired contractile efficiency and performance. Computational modelling showed that the ventricular contraction mechanism, structure, and electrophysiology underpin this behavior in healthy hearts and exacerbate it in disease.

**Conclusion:** Left ventricular systolic function is characterized by a myocardial tug-of-war at the mesoscale, which contributes to the heart’s adaptability in health and its vulnerability in disease. These findings introduce a new concept for understanding left ventricular function and a novel analytical approach for investigating its failure.

## Introduction

The contraction of the heart is dependent on the activity of more than two billion cardiomyocytes. These cells are arranged in an anisotropic fiber structure,^1,2^ and are activated by a sequential electrical depolarization wave.^3,4^ Between the microscale of individual cardiomyocytes and the macroscale of left ventricular regions, this structural and electrical organization forms up to millimeter-sized units of myocardium that operate at the intermediate, mesoscale level.^5,6^

Left ventricular function relies on the shortening of structurally and electrically coupled mesoscale units. As a result of this organization, mechanical non-uniformity is inherent to the normal function of the heart.^7–13^ Yet prior studies have predominantly explored non-uniformity at the macroscale, where it is largely interpreted as a pathological feature.^14–16^ At this level, ischemia and dyssynchrony create mechanical interactions resembling a “tug-of-war” between normally contracting and dyskinetic or dyssynchronous regions.^17–20^ In heart failure with intraventricular dyssynchrony, such large-scale interactions may worsen left ventricular function and patient outcomes if uncorrected.^21^ However, smaller-scale interactions between individual myocardial units remain largely unexplored. We hypothesize that the left ventricular contraction mechanism, structure and electrophysiology inherently generate a myocardial “tug-of-war” at the mesoscale, and that these interactions represent a previously unrecognized contributor to left ventricular function and failure.

To investigate this hypothesis, we employed a motion-encoded magnetic resonance imaging (MRI) technique that allows imaging of the beating heart with high temporal and spatial resolution.^22–24^ Using this technique, we comprehensively examined left ventricular function at millimeter resolution (i.e. the mesoscale level) and uncovered a distinct mechanical behavior (Fig. 1). We describe this mesoscale behavior as a myocardial tug-of-war, in which weakly contracting myocardial units are transiently elongated by the contraction of adjacent, more strongly contracting units. The aim of this study was to understand this interplay, its contribution to left ventricular function, and its plausible causes.

**Fig. 1.**
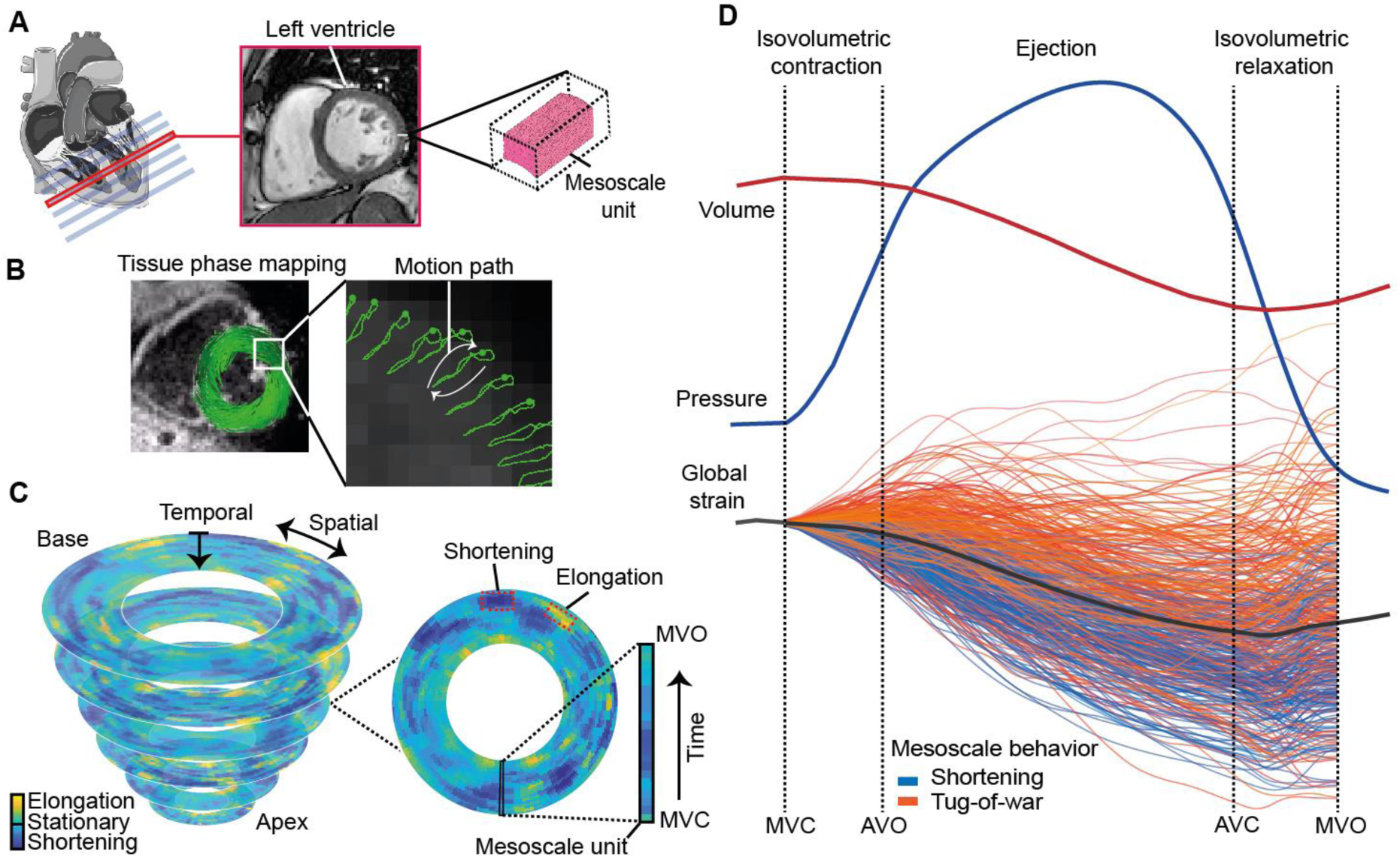
Mesoscale tug-of-war behavior in the heart. **a**, Diagram of the heart showing magnetic resonance imaging slices and a representative mesoscale unit. **b,** High-resolution tissue phase mapping image of the left ventricle illustrating tissue motion with representative motion paths (used to calculate strain) over the cardiac cycle. **c,** Mesoscale strain measured by tissue phase mapping from mitral valve closure (MVC, outer border) to mitral valve opening (MVO, inner border) in a representative heart. **d,** Wiggers diagram of the left ventricular volume, intraventricular pressure, global strain and mesoscale strain from the same subject as in c. Volume and strain data were obtained from same-session magnetic resonance imaging cine and tissue phase mapping, respectively. Mesoscale strain traces have been color coded to indicate uninterrupted contractions (simple shortening behavior) or complex tug-of-war behavior, containing transient phases of elongation. The intraventricular pressure curve is derived from a standard pressure curve (see methods) scaled to peak systolic brachial pressure and aligned to valvular events of the subject. For visualization, measurements have been scaled in time and space and smoothed. Raw data are shown in Supplemental Figure 1. Image credit for diagrams in a: Servier Medical Art. AVC, aortic valve closure; AVO, aortic valve opening.

## Methods

A detailed description of the materials and methods is provided in the Supplemental Methods section. All data supporting the findings of this study are available from the corresponding author upon reasonable request.

### Study population

Participants were enrolled at Oslo University Hospital, Ullevål between 2018 and 2025 as part of ongoing clinical studies.^25,26^

20 myocardial infarction patients between 18 and 80 years of age were enrolled six months after a first event ST-elevation myocardial infarction. Patients were excluded if they had a prior myocardial infarction, left ventricular bundle branch block, cardiogenic shock or resuscitated cardiac arrest, history of severe concomitant disease, or were receiving experimental treatment as part of clinical trials. Baseline characteristics are summarized in Table S1, and detailed inclusion and exclusion criteria are found elsewhere.^25^

Healthy individuals between 18 and 80 years of age were enrolled if they presented with good health. Individuals with a history of cardiovascular disease, or serious concomitant disease such as chronic infection, renal failure or cancer were excluded. The study included 20 healthy participants (study cohort) and 11 healthy participants performing a dynamic handgrip exercise (exercise cohort; see Supplemental Methods). In addition, 20 healthy participants were included as a replication cohort for tug-of-war analysis. Baseline characteristics for the study and exercise cohorts are summarized in Tables S1 and S2, and detailed inclusion and exclusion criteria are found elsewhere.^26^

All procedures were conducted in accordance with the Declaration of Helsinki and the International Conference on Harmonization Guidelines for Good Clinical Practice. All participants provided written informed consent prior to inclusion. The study was approved by the regional ethics committee (REK Sør-Øst 2016/1223 and REK Sør-Øst 2016/1961).

### Sample size

For the myocardial infarction cohort, sample size was determined by availability. This was an exploratory study and all eligible patients were included. For the age– and sex-matched healthy cohort, we included an equal number of participants and no formal power analysis was performed. For the exercise cohort, we expected the mean paired effect size and standard deviation to be half of that observed between healthy individuals and infarction patients (i.e. mean difference = 5%, SD = 5%). This required 10 subjects for paired analysis with a two sided significance level of 5% and a power of 80%. Based on availability and to account for uncertainty in the power calculation, we included 12 participants. No data were excluded from analysis in the myocardial infarction or healthy cohort. One participant from the exercise cohort was excluded because of procedural error during the experiment.

### Magnetic resonance imaging

All MRI was performed on a 1.5 Tesla Philips Ingenia System (Philips Healthcare, Best, The Netherlands). The full MRI protocol is detailed in the Supplemental Methods section.

Cardiac function was evaluated using MRI tissue phase mapping (TPM), which uses bipolar magnetic gradients to encode myocardial motion information directly into the signal phase of the image at the pixel level.^22,27^ This motion encoding enables quantification of myocardial deformation (strain) with high spatial and temporal resolution without reliance on tracking anatomical features or speckles.^22^ Nine short-axis TPM slices, each with a slice thickness of 10 mm, were acquired. We excluded the most apical slice to avoid interference from the longitudinal component of the myocardial wall in the apex and the two most basal slices to avoid interference from the aortic root, mitral valve ring, and associated structures. Six slices were therefore used for analysis. In-plane resolution ranged from 1.68×1.68 to 1.94×1.94 mm² (median 1.94×1.94 mm^2^). Echo time ranged from 3.66 to 4.48 ms (median 4.25 ms), and repetition time from 13.2 to 26.1 ms (median 20.0 ms). 50 frames per heart cycle were reconstructed from the data, and the flip angle was set to 10°. The velocity encoding strength (V_enc_) was set to 20 cm/s.

### Analysis of mesoscale function

TPM images were exported to MATLAB R2020b and segmented by an experienced observer. The only observer-dependent step was the use of a previously validated semi-automatic segmentation method with excellent reproducibility for motion-encoded strain analysis.^28^ For mesoscale analysis, circumferential strain was analyzed across transmural myocardial units spanning approximately 1-2 pixels (2.6 ± 0.4 mm) in width at the midmyocardium, using a validated post-processing framework for accurate high-resolution regional strain analysis.^29^ A full description is provided in the Supplemental Methods section.

To ensure that noise levels did not represent a systematic bias between units, velocity-to-noise ratio was calculated in each mesoscale unit. The velocity-to-noise ratio measures the signal strength of the velocity information compared to the noise present in the image,^30^ and was calculated using a previously validated method.^31^ A full description is found in the Supplemental Methods section.

Mechanical inefficiency was derived from measurements of myocardial work calculated from regional surface tension-strain loops, as previously described.^32^ Myocardial work measurements were then filtered into being wasted or constructive work.^33^ Wasted work was defined by systolic elongation (between mitral valve closure and aortic valve closure) and shortening during isovolumetric relaxation, while constructive work was defined as the inverse. Mechanical inefficiency was defined as the ratio of wasted work to the sum of wasted and constructive work.

### Analysis of tug-of-war behavior

#### Peak detection and definition

The strain trajectory in each mesoscale unit was tracked from the start of contraction (i.e. QRS in electrocardiogram) to the start of filling (i.e. mitral valve opening). Inclusion of the isovolumetric relaxation phase ensured that post-systolic shortening was captured. Transitory shortening and elongation phases were identified by peaks in the trajectory. The lower threshold for peak prominence was set to 0.5 % strain, as this effectively identified shortening and elongation phases. The threshold was determined by visual examination of the curves prior to any statistical analysis, and was later reproduced at a higher (1 %) and lower (0.25 %) strain thresholds. Peaks were then classified as upwardly oriented, Positive-(P)-peaks, or with downward orientation, Negative-(N)-peaks.

For each peak, we measured amplitude, timing, and in-slice spatial localization. Peak strain was defined as the peak with the largest negative value. Timing was measured in absolute units (ms) and relative to valvular events (scale 0-100), where 0-20 marked the isovolumetric contraction, 20-80 the ejection phase, and 80-100 the isovolumetric relaxation.

#### Contraction pattern classification

Strain curves exhibiting one or more P-peaks were classified as affected by tug-of-war behavior. Trajectories lacking P-peaks were classified as exhibiting simple shortening behavior. If the trajectory contained a P-peak before reaching 50% of the ejection interval, it was defined as exhibiting early tug-of-war behavior. If the trajectory contained a P-peak at or after reaching 50% of the ejection interval, it was classified as late tug-of-war behavior. The presence of tug-of-war in the heart was calculated as the proportion of strain curves with P-peaks. This value was calculated globally across all mesoscale units in the heart, and in specific regions of interest, providing a metric of regional tug-of-war behavior.

#### Spatiotemporal maps

In-slice localization of strain peaks was determined using spatiotemporal strain-rate maps, where the y-axis reports the circumferential (spatial) position and the x-axis is time. The temporal distribution was calculated as the average number of P– and N-peaks at each time point relative to valvular events. For determining the spatial proximity between in-slice P– and N-peaks, we considered peaks occurring simultaneously or within one time frame (20.5 ± 3.0 ms). The distance between each P-peak and the nearest N-peak was measured in mesoscale units and then converted to millimeters by multiplying by the units’ average sector width.

#### Penetration maps

Analysis was repeated at lower resolutions, aggregating mesoscale units by a factor of x2, x4, and x12. All analysis, including peak definition, contraction pattern analysis, peak magnitude, and myocardial inefficiency, was performed in an identical manner at all resolution levels.

### Computational modelling

To assess the types of force imbalances that could plausibly produce tug-of-war behavior, we used a computational model of cardiac excitation–contraction coupling. The model consisted of a ring of multiple contractile units as based on the framework of Rice et al.^34^ This allowed us to introduce variability in contractile unit resting length and/or Ca^2+^ handling, which we produced under time-dependent external loads that mimic isovolumetric contraction, ejection, and isovolumetric relaxation. A full description of the model is provided in the Supplemental Methods “Cardiac excitation-contraction modelling” section with parameter values found in Table S3.

To assess the plausible impact of left ventricular structure and electrophysiology on tug-of-war behavior, we employed a finite-element model of the left ventricle. Here the left ventricle was represented as a half-ellipsoid with configurable fiber structure and electrical activation sequence, which enabled us to change each of these components individually. Loading conditions were chosen to produce a physiologically realistic pressure–volume loop and were held constant across experiments. A full description of the model is provided in the Supplemental Methods “Finite element left ventricular modelling” section. The modelling pipeline is available as a Python package on GitHub: <u>GitHub – mjsadeghinia/TugOfWar</u>.

### Statistical analysis

Investigators were blinded to the heart’s identity during analysis. All statistical analysis were performed using GraphPad Prism version 10 (GraphPad Software Inc., San Diego, USA). Continuous data were summarized using mean and standard deviation unless otherwise noted. Normality was assessed with the Shapiro–Wilk test. Normally-distributed data sets were tested by Student’s t-tests, or ANOVA when two or more samples were compared. When assumptions of normality were violated, the Mann–Whitney test, Wilcoxon matched-pairs signed-rank test, or Kruskal–Wallis test was applied, as appropriate. Comparisons were made at the aggregated level, yielding one value per heart. Correlation was performed using Pearson’s test. A two-tailed P-value <0.05 was considered statistically significant.

## Results

### Tug-of-war is prevalent in health and increases in disease

Across the left ventricle, myocardial strain (deformation) was measured in 427 ± 16 millimeter-sized units per heart, revealing pronounced spatial non-uniformity at the mesoscale (Fig. 1ab). Indeed, during the contraction cycle of the heart between mitral valve closing and opening, many myocardial units exhibited phases of elongation (Fig. 1c). Thus, a complex *tug-of-war* pattern was apparent within these units, with periods of negative and positive strain (shortening and elongation), which distinguished them from units which purely shortened (Fig. 1d).

After confirming that non-uniformity between myocardial units did not reflect imaging acquisition noise (Supplemental Fig. 1), showed consistent features between healthy and infarcted hearts across varying peak thresholds (Supplemental Fig. 2), and replicated these contraction patterns in an independent healthy cohort (Supplemental Fig. 3), we examined mesoscale strain in greater detail (Fig. 2a). In units affected by tug-of-war behavior, we observed a bimodal distribution of elongation and shortening phases during the cardiac cycle (Fig. 2b). *Early* tug-of-war behavior occurred during isovolumetric contraction and the first half of ejection, reflecting alternating phases of shortening and elongation as the heart begins to contract. *Late* tug-of-war behavior occurred during the second half of ejection and isovolumetric relaxation, reflecting alternating phases of shortening and elongation as the heart begins to relax.

**Fig. 2.**
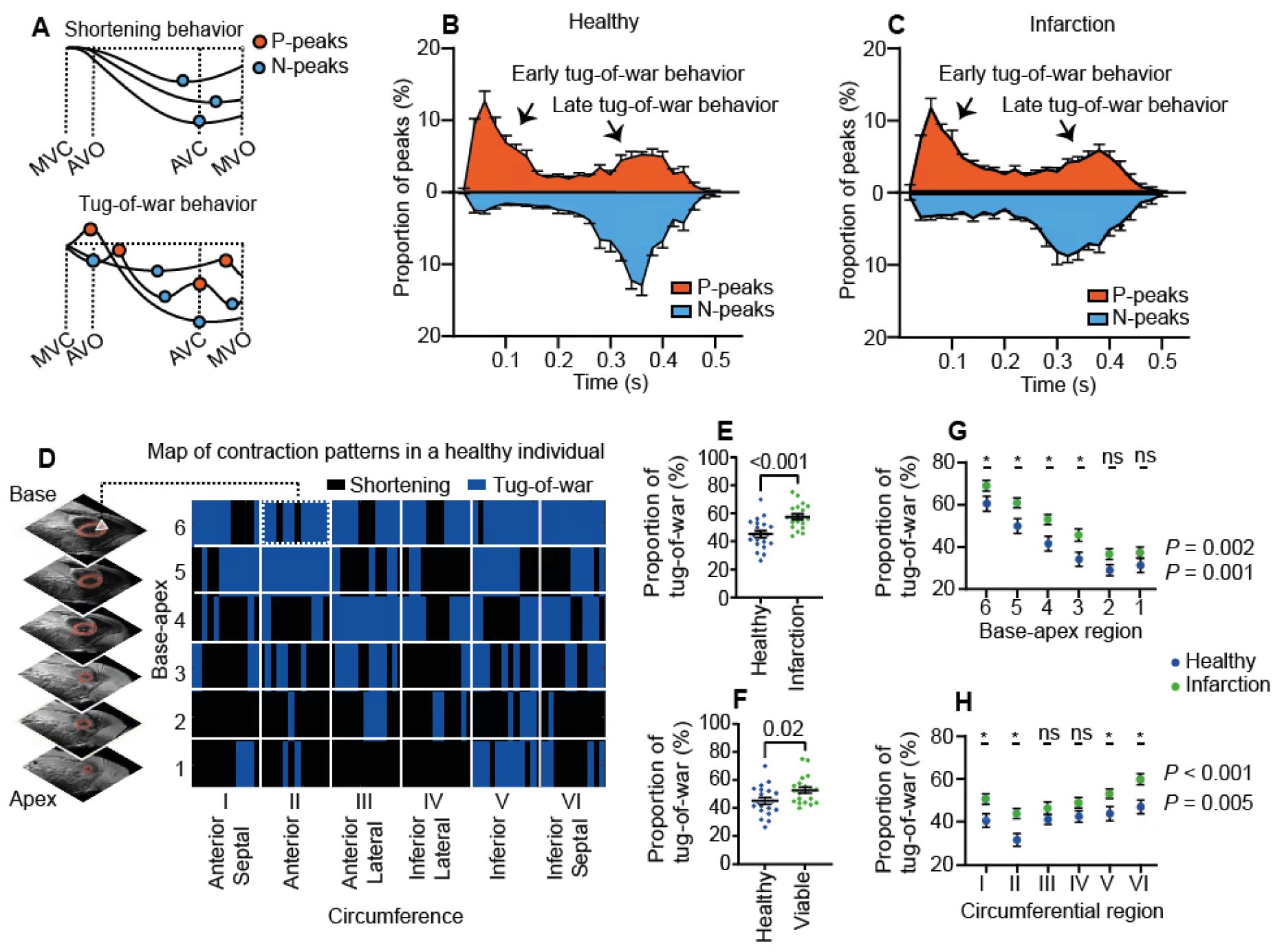
Tug-of-war varies across the left ventricle and increases after myocardial infarction. **a**, Diagram of mesoscale units exhibiting simple shortening and complex tug-of-war behavior. Positive (P) and negative (N) strain peaks are indicated. P-peaks mark the end of an elongation phase, whereas N-peaks mark the end of a shortening phase. Units affected by tug-of-war display phases of elongation and contain therefore, by definition, one or more P-peaks. Units exhibiting simple shortening display no elongation phase and contain therefore, by definition, only one N-peak. **b,c,** Frequency distributions of P– and N-peak timings, in healthy individuals (b) and patients with myocardial infarction (c). The first population of P-peaks reflects early tug-of-war behavior. The second P-peak population reflects late tug-of-war behavior. **d,** Map of mesoscale contraction pattern distributed across the left ventricle in a healthy individual. The left ventricle is divided into regions by circumference (columns) and from apex to base (rows). **e,f**, Comparison of tug-of-war behavior between healthy and infarcted hearts (e), and between healthy hearts and the non-infarcted, viable myocardium of infarcted hearts (f). Group comparisons using Student’s T-test for unpaired groups. **g,h,** Spatial distribution of regional tug-of-war presence from apex to base (g), and around the circumference (h) in healthy individuals and in patients with myocardial infarction. * *P* < 0.05 using Student’s T-test for unpaired groups. Reported *P* values are ANOVA test for linear trend. Data are mean ± standard error of the mean, with *n* = 20. AVC, aortic valve closure; AVO, Aortic valve opening; MVC, mitral valve closure; MVO, mitral valve opening.

Notably, the overall characteristics of tug-of-war behavior were similar in both healthy individuals and in patients with myocardial infarction (Fig. 2b,c) and were observed throughout the left ventricle (Fig. 2d). However, tug-of-war behavior was more prevalent in infarction patients, where it was observed in 58 ± 9 % of myocardial units compared to 45 ± 11 % of units in healthy volunteers (*P* < 0.001) (Fig. 2e), and remained elevated in the non-infarcted viable myocardium (52 ± 10 %, *P* = 0.02) (Fig. 2f). In both types of individuals, tug-of-war behavior was higher in basal compared with apical regions and in inferior compared to anterior regions, with consistently elevated levels in the infarcted heart (Fig. 2g,h).

### Tug-of-war reflects local mechanical interactions in the myocardium

Having characterized the non-uniformity of mesoscale strain across the heart, we next investigated its mechanical dependency. “Spatiotemporal maps” were constructed to determine both the spatial and temporal coordinates of transient elongation and shortening phases during the contraction cycle (Fig. 3a).

**Fig. 3.**
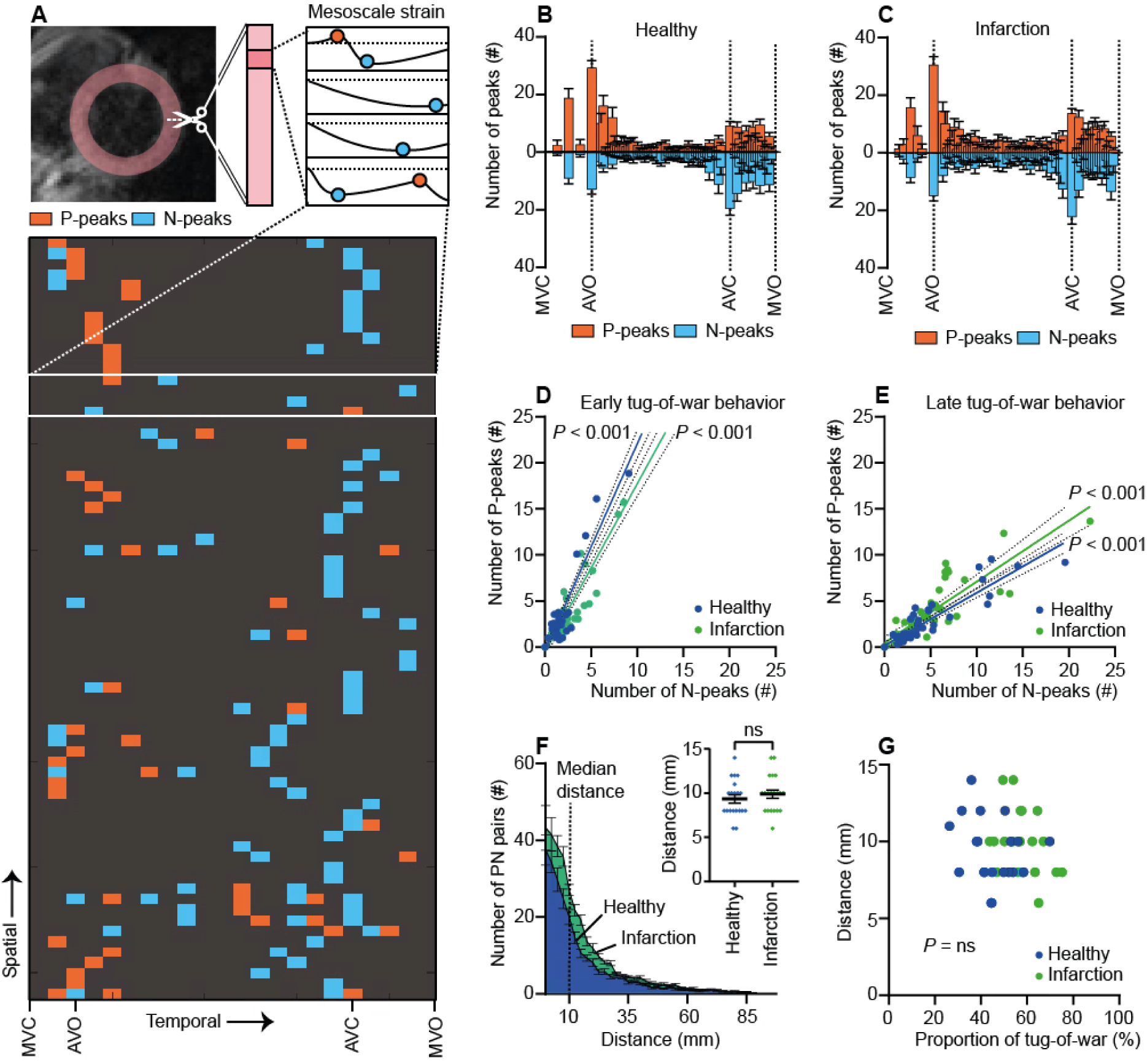
Spatiotemporal interactions within the left ventricle. **a**, Spatiotemporal map of P-and N-peaks from a representative individual, and a diagram of its construction process. The map shows the positioning of strain peaks in time (along the horizontal axis) and in space (along the vertical axis) within one imaging slice. **b,c,** Frequency distribution plots of P– and N-peak timing in units with tug-of-war behavior, shown relative to valvular events in healthy individuals (b) and in patients with myocardial infarction (c). **d,e,** Correlation between the number of P-peaks and N-peaks at each time point, shown separately for early (d) and late (e) tug-of-war behavior. **f**, Distribution of spatial distance between every P-peak and the nearest neighbor N-peak (PN-pairs). Top right: Comparison of median P– and N-peaks distances in healthy individuals and patients with myocardial infarction (*P* = 0.41, Student’s T-test). **g**, Correlation between median P– and N-peaks distance and the presence of tug-of-war in the heart, *n* = 40. Data are presented as mean ± standard error of the mean, with *n* = 20, unless otherwise noted. AVC, aortic valve closure; AVO, Aortic valve opening; MVC, mitral valve closure; MVO, mitral valve opening.

We first examined temporal interactions. Analysis of the timing of elongation and shortening events revealed that elongation in one myocardial unit consistently coincided with simultaneous shortening in another unit (Fig. 3b,c). Correlation analysis demonstrated this interdependency, revealing strong correlations between the elongation and shortening phases for every time point, both in units affected by early tug-of-war behavior (healthy: *r* = 0.97, *P* < 0.001; infarction: *r* = 0.96, *P* < 0.001), and in units affected by late tug-of-war behavior (healthy: *r* = 0.92, *P* < 0.001; infarction: *r* = 0.87, *P* < 0.001) (Fig. 3d,e).

Next, we examined spatial interactions, analyzing the nearest-neighbor distance between elongation and shortening phases. This analysis revealed that most transient elongation phases occurred in close spatial proximity to a transient shortening phase (Fig. 3f). This behavior was most probable between immediately neighboring units and decreased exponentially with increasing distance; a finding which was similar in healthy individuals and in patients with myocardial infarction (median distance, healthy and infarction: 10 ± 2.5 mm, *P* = 0.41) (Fig. 3f), and independent of the total presence of tug-of-war behavior in the left ventricle (median distance vs presence of tug-of-war: *r* = –0.16, *P* = 0.33) (Fig. 3g).

### Tug-of-war affects dysfunctional units

We observed that units exhibiting tug-of-war contracted less (peak strain) than units exhibiting simple shortening behavior (–13 ± 2 % vs –20 ± 2 %, *P* < 0.001) (Fig. 4a), and less in patients with myocardial infarction than in healthy individuals (–12 ± 2 % vs –14 ± 1 %, *P* = 0.001). These units also reached peak contraction (time to peak strain) earlier than shortening units (314 ± 35 ms vs 351 ± 32 ms, P < 0.001), indicating less sustained contraction during systole (Fig. 4b). Shortening units, on the other hand, displayed strong contractions that were similar between healthy individuals (–21 ± 2 %) and patients with myocardial infarction, both in magnitude (–20 ± 2 %, *P* = 0.3) and timing (healthy: 353 ± 32 ms; infarction: 348 ± 32 ms, *P* = 0.6) (Fig. 4a, b).

**Fig. 4.**
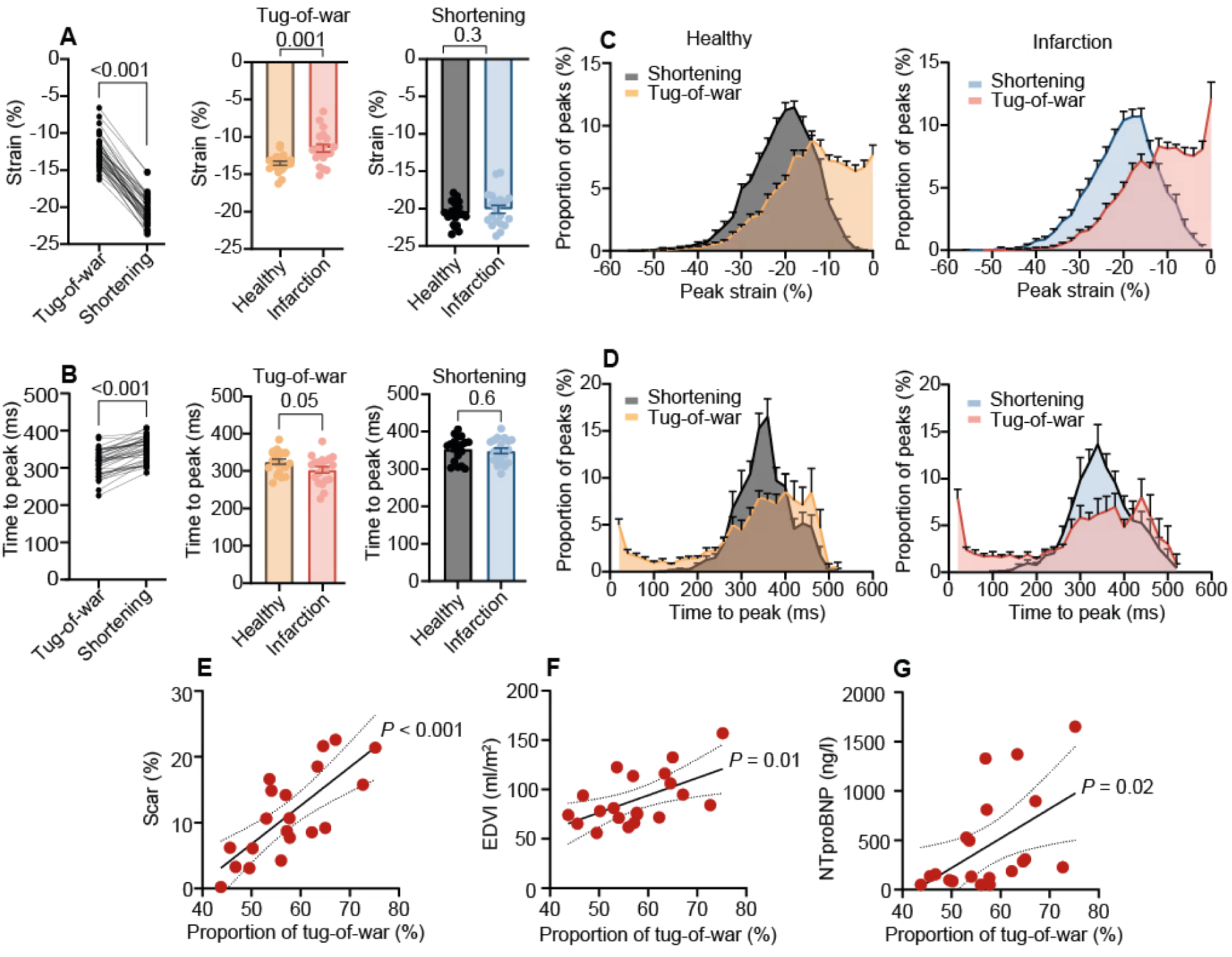
Tug-of-war affects dysfunctional units and increases with disease burden. **a,b**, Peak strain (a) and time-to-peak strain (b) in mesoscale units exhibiting tug-of-war and simple shortening behavior. **c,d**, Frequency distributions of peak strain (c) and time-to-peak strain (d) in units exhibiting tug-of-war and simple shortening behavior. **e-g**, Correlations between the presence of tug-of-war, infarct size (e) and indices of cardiac remodelling (f,g). Comparisons using paired (left panel a and b, *n* = 40) or unpaired Student’s T-test. Data are mean ± standard error of the mean, with *n* = 20 unless otherwise noted.

But does this difference between weakly and strongly contracting units account for the complex tug-of-war behavior observed? To investigate this question, we examined the distributions of contraction magnitudes and time courses. For those units exhibiting tug-of-war, both peak strain and timing of peak strain showed wide distributions with less coordinated contraction magnitudes and time courses in both healthy and myocardial infarction hearts (Fig. 4c,d). In contrast, units exhibiting shortening behavior displayed rather narrow distributions, reflecting more coordinated contractions (Fig. 4c,d).

The prevalence of tug-of-war behavior increased with greater infarct size (*r* = 0.75, *P* < 0.001) and the degree of post-infarction remodeling, as measured by end-diastolic volume index (*r* = 0.57, *P* = 0.01) and levels of N-terminal pro-B-type natriuretic peptide (NT-proBNP) (*r* = 0.52, *P* = 0.02) (Fig. 4e-g).

### Tug-of-war shapes left ventricular efficiency and performance

To understand the impact of tug-of-war behavior on systolic dysfunction, we first quantified the mechanical inefficiency arising from this behavior as the ratio of wasted work (work performed during systolic elongation or post-systolic shortening) to the total work produced by the myocardium (Fig. 5a).^33^ A strong correlation between the presence of tug-of-war behavior and inefficiency was observed (*r* = 0.79, *P* < 0.001), consistent with greater inefficiency in patients with myocardial infarction (19 ± 7 %) compared to healthy individuals (12 ± 4 %, *P* < 0.001) (Fig. 5b,c).

**Fig. 5.**
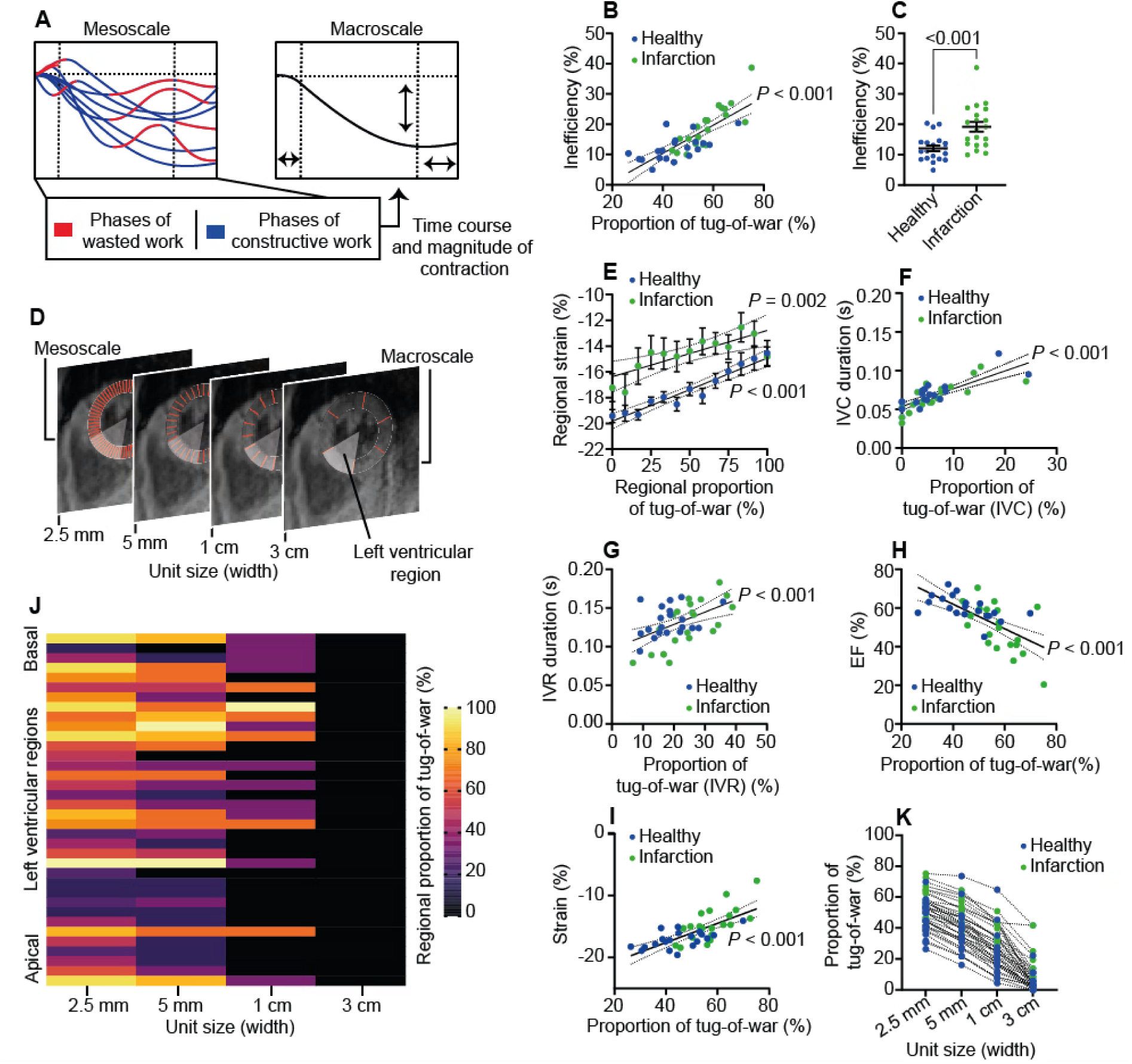
Tug-of-war shapes left ventricular efficiency and function. **a**, Diagram illustrating typical contraction patterns at the meso– and macroscale. At the mesoscale, the tug-of-war behavior creates phases of wasted and constructive work that influence time course and magnitude of contraction at the macroscale. **b**, Correlation between mesoscale tug-of-war and inefficiency in the left ventricle. Inefficiency is defined as the wasted work divided by the total work produced (wasted plus constructive work). **c**, Comparison of inefficiency between healthy and infarction hearts, *n* = 20. **d,** Diagram showing aggregation of mesoscale units into larger myocardial units, ultimately forming a macroscopic region in the left ventricle. **e,** Correlation between the proportion of mesoscale units exhibiting tug-of-war behavior within a macroscopic region and the contraction of the macroscopic region as a whole. The regions have been filtered by their underlying tug-of-war presence, from a minimum of 0% to a maximum of 100%, and are shown separately for healthy and infarction hearts, *n* = 20. **f,g,** Correlations between mesoscale tug-of-war presence during isovolumetric contraction (IVC) (f) and relaxation (IVR) (g), and the duration of those phases. **h,i,** Correlations between mesoscale tug-of-war presence and ejection fraction (EF) (h) and global strain (i). **j,** Representative heatmap of the regional tug-of-war presence across the heart shown as a function of increasing unit size, from the meso– to the macroscale. **k,** Mean tug-of-war presence across scales. Data are mean ± standard error of the mean, with *n* = 40 unless otherwise noted. Group comparisons using unpaired Student’s T-test.

But does greater tug-of-war and inefficiency reflect reduced left ventricular function? To investigate this question, we aggregated neighboring myocardial units into left ventricular macroscopic regions similar to the American Heart Association segmentation model (Fig. 5d).^35^ When we next measured the contraction of each macroscopic region, we observed a strong correlation between the underlying presence of tug-of-war and the contraction of the region as a whole, both in healthy individuals ( *r* = 0.97, *P* < 0.001) and patients with myocardial infarction (*r* = 0.78, *P* = 0.002) (Fig. 5e). Notably, the presence of tug-of-war also correlated with the duration of the isovolumetric contraction phase (*r* = 0.79, *P* < 0.001) and the duration of the isovolumetric relaxation phase (*r* = 0.51, *P* < 0.001) (Fig. 5f,g), as well as with ejection fraction (EF) (*r* = –0.65, *P* < 0.001) and global strain (*r* = 0.71, *P* < 0.001) (Fig. 5h,i).

### Tug-of-war is attenuated at the macroscale

An interesting phenomenon was observed when mesoscale myocardial units were integrated over a range of size of units. As shown by “penetration maps” (Fig. 5j), we found that visible tug-of-war behavior gradually diminished as the unit size increased (Fig. 5k), independent of the underlying presence of tug-of-war (Supplemental Fig. 4a). Consequently, only a median of 3 % (interquartile range: 0-5 %) of left ventricular macroscopic regions in healthy individuals displayed visible tug-of-war behavior (Supplemental Fig. 4b), mainly in the left ventricular base (Supplemental Fig. 4c). Comparable effects were observed in patients with myocardial infarction (Supplemental Fig. 4a), albeit with elevated macroscale levels of tug-of-war behavior in the area of infarction (infarct area median: 11 % [interquartile range 5-24 %], viable myocardium median: 3 % [interquartile range 0-9 %]) (Supplemental Fig. 4d), likely reflecting a greater force-imbalance in this area.

Next, we investigated whether the macroscopically diminishing tug-of-war behavior resulted from the cancellation of opposing forces between underlying myocardial units. We used spatiotemporal maps to identify whether neighboring units displayed simultaneous counteracting movements. Then, to assess cancellation or “smoothing” effects across the aggregated area, we combined this information with penetration maps. Through this approach we observed that smoothing occurred more often between counteracting myocardial units than between non-counteracting units (Supplemental Fig. 5a,b). Furthermore, when several myocardial units were aggregated, the influence of counteracting units extended beyond immediate neighbors to include the surrounding units as well (Supplemental Fig. 5c-g).

### Baseline levels of tug-of-war supports cardiac reserve capacity in the healthy heart

Having described the (patho)physiological role of tug-of-war behavior, we next sought to understand why it pervades the healthy heart. We hypothesized that baseline levels of tug-of-war might provide a capacity for cardiac reserve during times of acute physiological stress. In healthy individuals, performing a dynamic handgrip exercise moderately increased both global function (peak strain rest: –18 ± 3 %; stress: –19 ± 2 %, *P* = 0.04) and systolic blood pressure (rest: 119 ± 23 mmHg; stress: 136 ± 25 mmHg, *P* = 0.04) (Fig. 6a-c). Simultaneously, the proportion of myocardial units affected by tug-of-war behavior decreased (rest: 53 ± 10 %; stress: 48 ± 11 %, *P* = 0.03) (Fig. 6d), suggesting a more uniform and potentially more efficient myocardial performance under acute stress. The remaining myocardial units affected by tug-of-war behavior displayed similar early and late features (Fig. 6e).

**Fig. 6.**
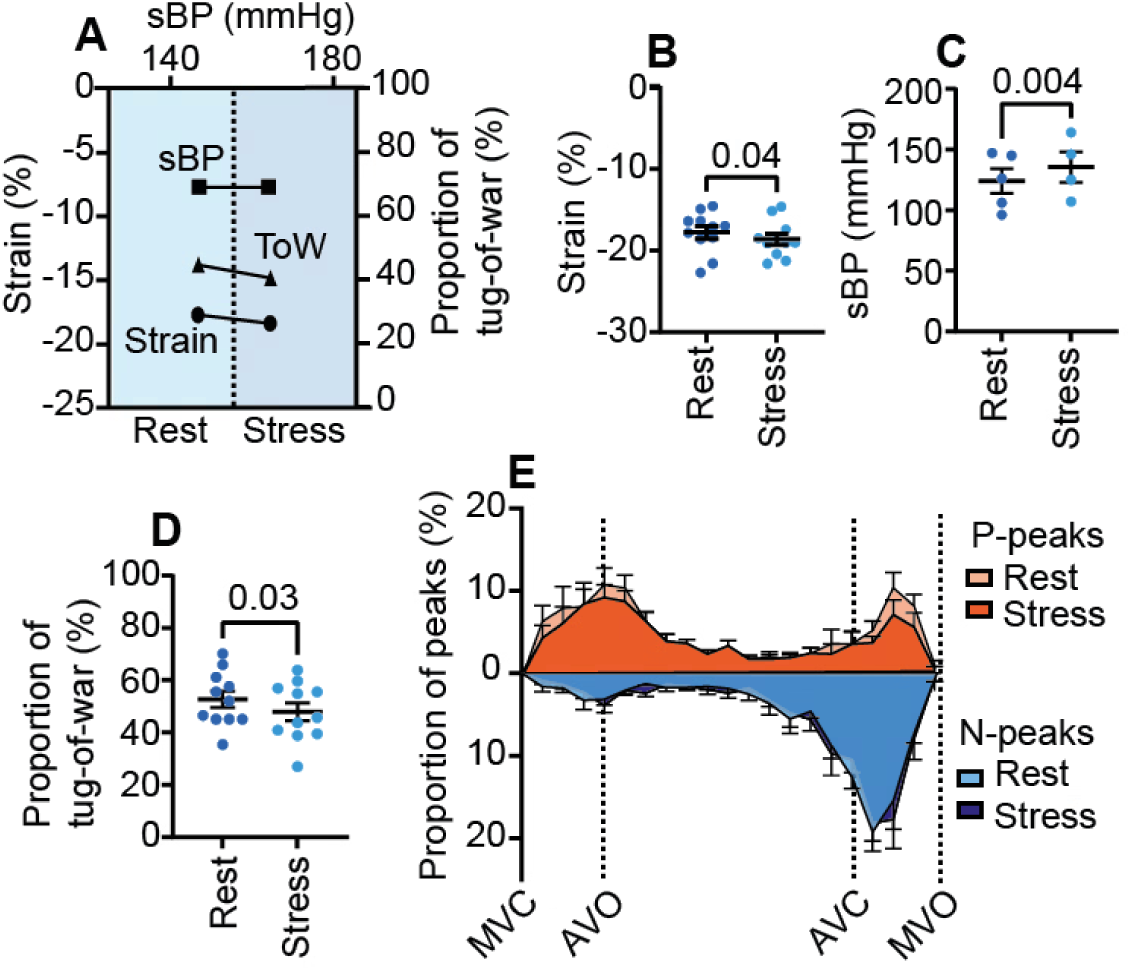
Baseline levels of tug-of-war and cardiac reserve capacity. **a**, The response to acute stress (dynamic handgrip exercise) in a representative healthy subject. **b-d,** Global strain (b), systolic blood pressure (sBP) (c) and global tug-of-war presence (d) at rest and stress. **e,** Frequency distribution plots of P– and N-peak timings in mesoscale units at rest and stress. Data are mean ± standard error of the mean, with *n* = 11. Comparisons using paired Student’s T-test. AVC, aortic valve closure; AVO, Aortic valve opening; MVC, mitral valve closure; MVO, mitral valve opening; ToW, tug-of-war.

### Contraction mechanism origins of tug-of-war

Having described the tug-of-war behavior in health and disease, we next investigated possible underlying causes. To this end, we employed a computational model of the cardiac contraction mechanism which included a description of excitation-contraction coupling.^34^ The model comprised a ring of multiple contractile units, analogous to myocardial units, with the possibility to introduce variability in contractile unit resting length or Ca²⁺-handling (Fig. 7a), as summarized in Table S3. This enabled us to assess the types of force imbalances within the myocardium that could plausibly generate tug-of-war behavior.

**Fig. 7.**
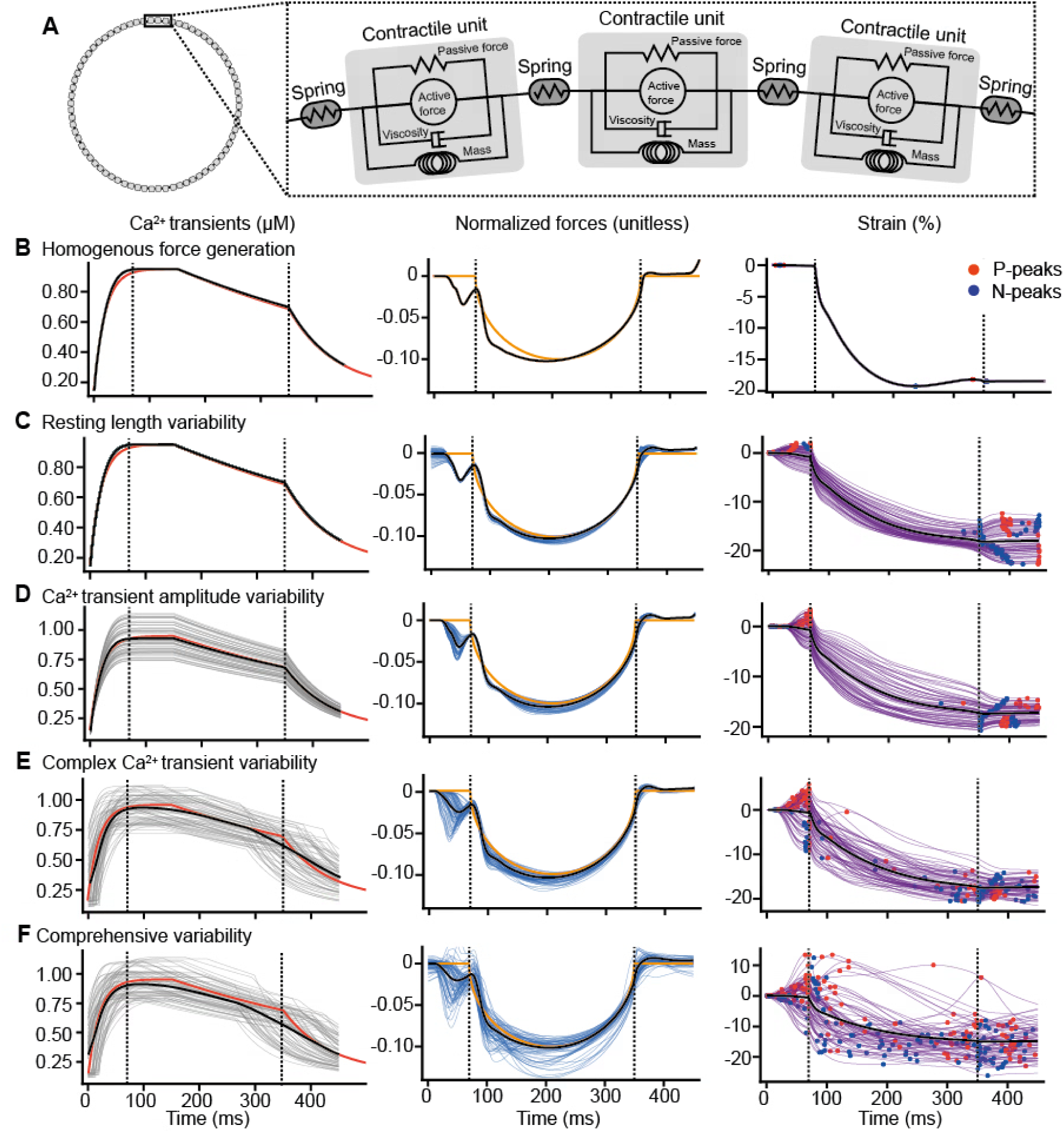
*In silico* tug-of-war behavior and the contraction mechanism of the heart. **a**, Diagram of the model. **b,** Ca²⁺ transients, forces and strains in baseline simulation with no variability. **c-e,** Ca²⁺ transients, forces and strains in simulations with variability in various components of the *in silico* model: end-diastolic length (resting length) (c); Ca²⁺ release magnitude (d); Ca²⁺ handling magnitude, influx/efflux rate and de/repolarization timing (e). **f**, Comprehensive simulation with variability in all components: contractile unit resting length, Ca²⁺-handling, and Ca²⁺-affinity. Thick black curves in all panels represent averages. The red curves in Ca^2+^ panels show the baseline Ca^2+^ profile. The orange curves in the force panels, shows the imposed external load.

First, we simulated homogeneous force generation, with no variability in contractile unit resting length or Ca²⁺-handling. Under these conditions we observed uniform contractions and no tug-of-war (Fig. 7b). When we next introduced variability in the end-diastolic length of the contractile unit (resting length),^36^ a tug-of-war behavior was observed, with the appearance of P– and N-peaks both early and late in the contraction cycle. Here, weaker units with shorter resting lengths were observed to be stretched by the stronger units with longer resting lengths (Fig. 7c).

When we introduced variability in Ca²⁺-transient amplitude,^37^ similar effects on tug-of-war behavior were observed (Fig. 7d). When simulating the effects of more complex Ca²⁺-handling variability, including differences in both Ca²⁺ release magnitude and time course as well as differences in myofilament Ca^2+^ sensitivity,^38–42^ the tug-of-war behavior was amplified (Fig. 7e). The individual effects of these alterations are summarized in Supplemental Fig. 6.

Lastly, we performed a comprehensive simulation, including summated variability in contractile unit end-diastolic length, Ca²⁺-handling transients, myofilament sensitivity, and the timing of depolarization and repolarization (Fig. 7f). This resulted in early and late tug-of-war behavior visually similar to our observations *in vivo*.

### Influence of left ventricular structure and electrophysiology on tug-of-war

Having identified potential aspects of the contraction mechanism that promote tug-of-war, we finally investigated whether left ventricular structure and electrophysiology might contribute. To this end, we used a finite element computational model incorporating both the fiber structure and electrophysiology of the left ventricle. Units were embedded in the model with dimensions and spatial distributions matching the in vivo data. We then analyzed contraction patterns in these units under different structural and electrophysiological conditions.

In a simplified and non-physiological simulation, assuming a homogenous fiber structure and synchronous electrical activation pattern, we observed uniform contractions with no tug-of-war (Fig. 8a). This configuration also yielded suboptimal systolic function, with an EF of 47%. When we introduced a physiological fiber structure, incorporating a transmural right to left-handed helix from the endocardium to the epicardial surface,^1,2^ we observed tug-of-war in 12% of the myocardial units (Fig. 8b). Notably, this structural change also improved the global performance, elevating EF to 58%. When we next added a physiological electrical activation pattern, sequentially propagating throughout the ventricle from apex to base and from the endocardium to the epicardium,^3^ we observed a pronounced effect elevating tug-of-war behavior to 58% while EF remained normal at 57% (Fig. 8c).

**Fig. 8.**
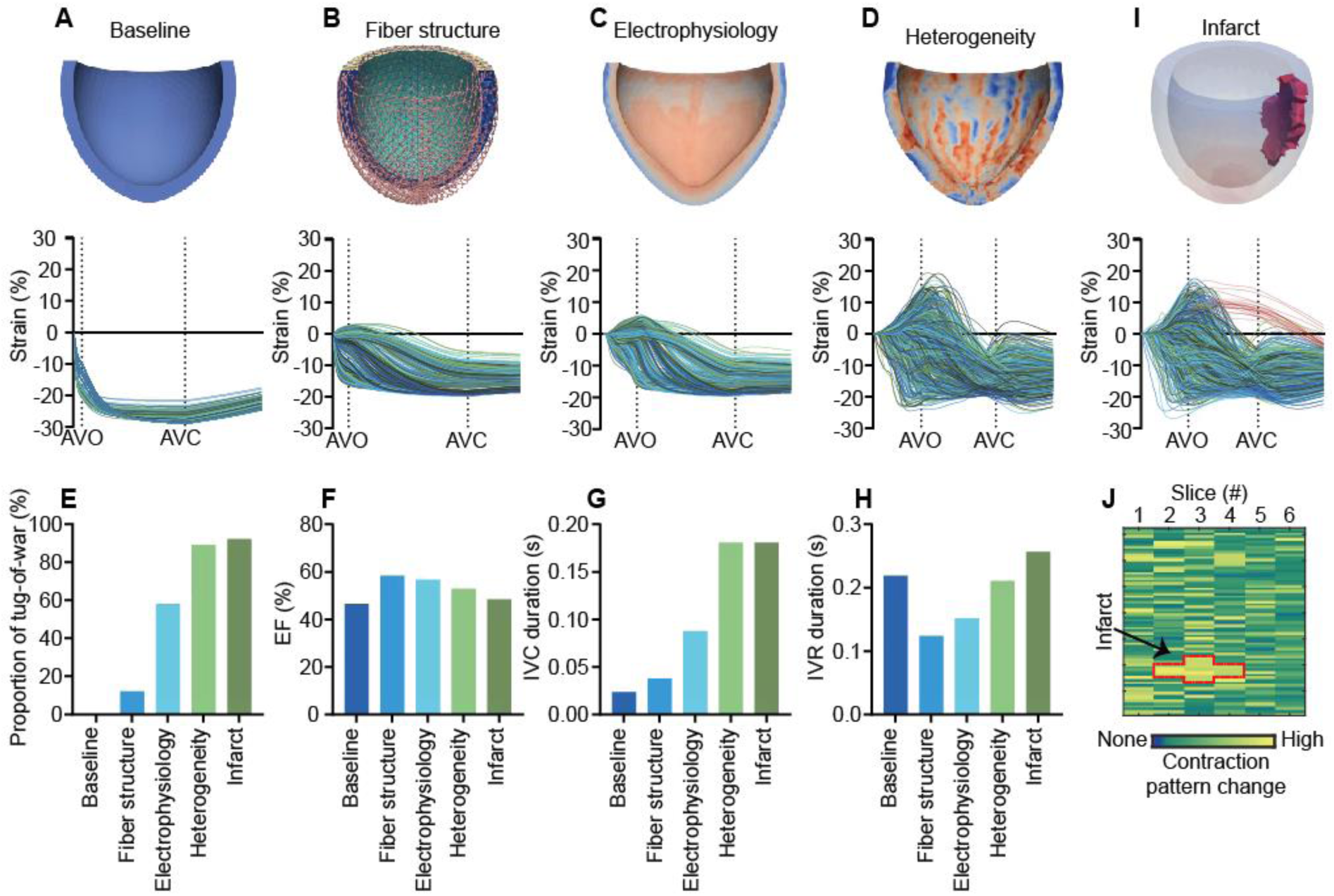
Left ventricular structure and electrophysiology generate tug-of-war *in silico*. **a**, Baseline simulation with a homogenous fiber structure and instantaneous electrical activation, and the resulting mesoscale contraction patterns. **b,** Simulation with a physiological fiber structure, consisting of an anisotropic transmural right– to left-handed helix from the endocardium to the epicardial surface. **c,** Simulation with fiber structure anisotropy and sequential electromechanical activation from apex to base and from the endocardium to the epicardium. **d,** Simulation including local discontinuities in activation timing, reflecting myocardial tissue heterogeneity arising from microvasculature, connective tissue, or trabeculation. **e-h**, Tug-of-war presence (e), ejection fraction (EF) (f) and duration of isovolumetric contraction (IVC) (g) and isovolumetric relaxation (IVR) (h) for the different simulations. **i**, Simulation with infarct and the resulting mesoscale contraction pattern, showing passive stretching in infarcted mesoscale units (red strain curves). **j**, “Change map” showing alterations in mesoscale contraction patterns when simulating with infarct compared to the simulation without infarct.

To examine the sensitivity to myocardial tissue heterogeneity, such as might result from pathological changes in microvasculature, connective tissue, or trabeculation,^6^ we randomly introduced local discontinuities in the electrical propagation wave (Fig. 8d). This generated markedly elevated tug-of-war (89%) and an EF of 53%. The effects of altering the gross structure and electrophysiology of the left ventricle on global function and the time course of the heartbeat is summarized in Figure 8e-h.

Finally, we introduced a local perturbation in the model, simulating a non-contractile “infarcted” area in the myocardium (Fig. 8i). This perturbation appeared to create a “domino effect” of force imbalances across the left ventricle, increasing tug-of-war to 92% while altering mechanical interactions throughout the viable myocardium (Fig. 8j).

## Discussion

Using high-resolution MRI and advanced post-processing methods, we show that left ventricular systolic function is influenced not only by the contraction of each myocardial unit but also by how it mechanically interacts with neighboring mesoscale units. Through computational modelling we suggest that these interactions can arise from heterogeneity in the contraction mechanism, structure, and electrophysiology of the left ventricle. This creates local force imbalances within the myocardium, generating a tug-of-war between myocardial units. These counteracting, inefficient contractions effectively cancel one another out, making the tug-of-war phenomenon hidden with lower resolution imaging approaches. We nevertheless show that tug-of-war affects contractile efficiency, presenting a basis for cardiac reserve in the healthy heart but impaired left ventricular performance in the diseased heart.

Given the intrinsic contractile, structural and electrical complexity of the myocardium, regional non-uniformity in contraction is perhaps inescapable.^43^ Indeed, non-uniformity has long been proposed as a central feature of both left ventricular function and failure.^7^ However, most imaging studies have primarily focused on the mechanics of centimeter-sized (macroscale) left ventricular regions, such as those defined by the American Heart Association segmentation model.^44^ At this level, mechanical non-uniformity is often interpreted as a pathological feature, such as early-systolic lengthening and post-systolic shortening during ischemia, or septal flash in left bundle branch block.^45–52^ These macroscale phenomena likely reflect mechanical interactions between normally contracting regions of the ventricle and those that are dyskinetic or dyssynchronously contracting.^17–20^ At the mesoscale, our findings reveal strain patterns resembling early-systolic lengthening and post-systolic shortening. In both healthy and diseased hearts, this tug-of-war is broadly distributed across the myocardium and not confined to specific pathological perturbations. This pervasiveness likely reflects the complex structure-function relationship of the myocardium, as supported by our computational modeling data.

The myocardial fiber structure and sequential activation pattern are known to generate transient patterns of shortening and elongation during the isovolumetric phases.^9–11^ By comprehensively examining these contraction patterns at the mesoscale, we confirm and build upon these observations. Our results show that mechanical interactions are closely related to contractile efficiency but become undetectable when they are aggregated over larger left ventricular regions (macroscale). This observation implies that contractility does not scale directly from cardiomyocytes (microscale) to global pumping function (macroscale), but that it is also shaped by mechanical interactions at the mesoscale. We therefore propose that mesoscale tug-of-war represents a distinct contractile phenomenon and a previously unrecognized determinant of left ventricular function.

Central to cardiac function is adaptability, which allows continuous response to changing demands of oxygen and nutrients.^53^ In physiological systems, adaptability is often ensured through resting conditions that enable reserve capacity to be recruited under stress.^54^ In the heart, this adaptability includes compensatory responses to increases in preload, afterload, sympathetic tone, and heart rate.^55^ Here, there are well-described cellular mechanisms that contribute to the compensatory response, including length-dependent activation and the cardiomyocyte’s response to increasing pacing frequency and β-adrenergic stimulation.^56–59^ The handgrip exercise performed in our study likely draws on all of these aspects to increase left ventricular performance. However, our results suggest the possibility of a larger scale contribution to cardiac reserve, as functional units of myocardium better coordinate their contractions, leading to declining tug-of-war behavior and improved left ventricular efficiency at the mesoscale. Future work should be aimed at dissecting the individual effects of preload, afterload, β-adrenergic tone, and heart rate on myocardial tug-of-war to more fully understand the quantitative contribution of this mechanism to cardiac reserve, and how this may change in disease. In this context, investigating how augmented tug-of-war contributes to the hallmark reduction in cardiac reserve (exercise intolerance) that occurs during heart failure represents a topic of particular interest.

In disease, cellular, structural, and electrical heterogeneity across the heart can become excessive.^60–62^ Our findings suggest mechanical consequences of this pathological heterogeneity, whereby an increased number of mesoscale units are engaged in tug-of-war behavior. Left ventricular dysfunction is therefore consistent with a complex mechanical interplay that involves both weaker contractions (i.e. reduced peak strain) and elevated levels of wasted mechanical work. We speculate that this mechanical inefficiency contribute to progressive dysfunction, compromising the heart’s energy reserves and accelerating its transition to overt heart failure.^63^ Furthermore, therapeutic interventions that reduce tug-of-war, e.g., via reducing Ca²⁺-handling variability or improving electrical activation patterns, could perhaps influence left ventricular function independently of direct changes in cellular contractility itself. Future studies are needed to determine whether tug-of-war behavior represents a therapeutically modifiable driver of dysfunction.

The precise mechanisms underlying tug-of-war are intriguing. At the microscale, a tug-of-war emerges between sarcomeres,^64,65^ as shorter sarcomeres are stretched by their elongated, stronger neighbors.^66^ At the macroscale, a tug-of-war emerges between dyskinetic and viable myocardium during ischemia,^15^ and between the septum and the left ventricular free wall during left bundle branch block.^18,19^ Although this behavior appears phenomenologically similar across scales, the underlying mechanisms likely differ. At the mesoscale, our modelling data suggest that local differences in the contraction mechanism, including differences in length dependent activation or Ca^2+^ transient parameters,^36–42^ can sufficiently vary force production to stretch weaker regions by their more forceful neighbors. However, our data additionally suggest that tug-of-war is dependent on the gross structure and electrophysiology of the left ventricle. Here, a surprising finding was that including left ventricular anisotropic structure and sequential electrical activation pattern simultaneously augmented tug-of-war *and* ejection fraction. This observation suggests that given the complex organization of the myocardium, tug-of-war may not only be inevitable but also allow fine-tuned contraction of the left ventricle as a whole. In the diseased heart, sources of excessive myocardial heterogeneity, such as fiber disarray,^67^ fibrosis,^61^ microvasculature,^68^ or altered cellularity,^69^ may compromise these benefits; a point which warrants further investigations.

This study has limitations. Our motion-encoded MRI technique measures myocardial strain at near-pixel resolution but remains specialized and not widely implemented. The analytical framework itself, however, is in principle modality-independent and could be adapted to more accessible imaging techniques. Larger and more diverse cohorts are warranted to confirm generalizability, and a more comprehensive, graded stress protocol would likely elicit greater changes in strain and tug-of-war dynamics. Our human data are also observational, and the causal role of tug-of-war in left ventricular dysfunction cannot be established by association alone. However, our computational experiments impose heterogeneity as an independent variable and reproduce tug-of-war as a direct mechanical consequence, and the handgrip manipulation, although modest in effect size, produced coherent changes in tug-of-war alongside global ventricular performance. Moreover, the principle that force imbalance between coupled units can affect performance is supported by observations at the microscale in sarcomeres and at the macroscale in left bundle branch block.^21,66^ Finally, the structure–function relationship of the myocardium is more complex than can be fully captured by millimeter-resolution imaging or by current computational models. Defining how electrical, structural, and contractile heterogeneity integrate across scales in health and disease would be an important direction for future work.

In conclusion, our findings reflect that left ventricular systolic function is inherently inefficient at the mesoscale. Influenced by the contraction mechanism, structure, and electrophysiology of the left ventricle, the resulting tug-of-war contributes to the heart’s adaptability in health and its vulnerability in disease. We therefore propose a new concept for understanding left ventricular function, along with an analytical approach for investigating its failure.

## Data Availability

All data supporting the findings of this study are available from the corresponding author upon reasonable request.

## Acknowledgements

M.B.H. conceived the concepts and was involved in data collection, analysis, interpretation and drafting the manuscript. I.S. and E.K.S.E. supervised the project and interpreted data, with E.K.S.E. also responsible for post-processing imaging data. Y.D.M.R. conducted stress experiments. R.S. was responsible for excitation-contraction modelling. M.J.S., S.W., J.S. were responsible for finite element modelling. J.A.B., K.B., G.Ø.A. were responsible for coordinating participant recruitment. N.E.K. was responsible for coordinating the imaging examination. V.R. was responsible for imaging data collection. T.H.S. helped establish imaging methodology. K.B.B. recruited healthy participants. G.S., M.S., W.E.L., J.L. interpreted data. All authors performed critical review of the manuscript and approved the final version.

The authors would like to thank Kaja Knudsen Bergo for helping including participants, and Anne Britt Woll, Mary Bolstad, Sofia H. Darmo Ovarsson, Siri Kristine Søisdal, Mads Melvold, Bente Elisabeth Østbøll Flaen and Wenche Synnøve Andreassen for imaging data collection.

## Abbreviations

EF: Ejection fraction
LGE: Late gadolinium enhancement
MRI: Magnetic resonance imaging
N-peak: Negative peak
NT-proBNP: N-terminal pro-B-type natriuretic peptide
P-peak: Positive peak
TPM: Tissue phase mapping

## References

1. Greenbaum RA, Ho SY, Gibson DG, Becker AE, Anderson RH. Left ventricular fibre architecture in man. Br Heart J. 1981;45:248–263. doi: 10.1136/hrt.45.3.248

2. Streeter Jr DD, Spotnitz HM, Patel DP, Ross Jr J, Sonnenblick EH. Fiber orientation in the canine left ventricle during diastole and systole. Circulation research. 1969;24:339–347.

3. Durrer D, van Dam RT, Freud GE, Janse MJ, Meijler FL, Arzbaecher RC. Total excitation of the isolated human heart. Circulation. 1970;41:899–912. doi: 10.1161/01.cir.41.6.899

4. Ramanathan C, Ghanem RN, Jia P, Ryu K, Rudy Y. Noninvasive electrocardiographic imaging for cardiac electrophysiology and arrhythmia. Nature Medicine. 2004;10:422–428. doi: 10.1038/nm1011

5. Wilson AJ, Sands GB, LeGrice IJ, Young AA, Ennis DB. Myocardial mesostructure and mesofunction. Am J Physiol Heart Circ Physiol. 2022;323:H257–h275. doi: 10.1152/ajpheart.00059.2022

6. Kléber AG, Rudy Y. Basic Mechanisms of Cardiac Impulse Propagation and Associated Arrhythmias. Physiological Reviews. 2004;84:431–488. doi: 10.1152/physrev.00025.2003

7. Brutsaert DL. Nonuniformity: a physiologic modulator of contraction and relaxation of the normal heart. J Am Coll Cardiol. 1987;9:341–348. doi: 10.1016/s0735-1097(87)80387-x

8. Voigt JU, Lindenmeier G, Exner B, Regenfus M, Werner D, Reulbach U, Nixdorff U, Flachskampf FA, Daniel WG. Incidence and characteristics of segmental postsystolic longitudinal shortening in normal, acutely ischemic, and scarred myocardium. J Am Soc Echocardiogr. 2003;16:415–423. doi: 10.1016/s0894-7317(03)00111-1

9. Sengupta PP, Korinek J, Belohlavek M, Narula J, Vannan MA, Jahangir A, Khandheria BK. Left ventricular structure and function: basic science for cardiac imaging. J Am Coll Cardiol. 2006;48:1988–2001. doi: 10.1016/j.jacc.2006.08.030

10. Sengupta PP, Khandheria BK, Korinek J, Wang J, Jahangir A, Seward JB, Belohlavek M. Apex-to-Base Dispersion in Regional Timing of Left Ventricular Shortening and Lengthening. Journal of the American College of Cardiology. 2006;47:163–172. doi: doi:10.1016/j.jacc.2005.08.073

11. Sengupta PP, Khandheria BK, Korinek J, Wang J, Belohlavek M. Biphasic tissue Doppler waveforms during isovolumic phases are associated with asynchronous deformation of subendocardial and subepicardial layers. Journal of Applied Physiology. 2005;99:1104–1111. doi: 10.1152/japplphysiol.00191.2005

12. Bogaert J, Rademakers FE. Regional nonuniformity of normal adult human left ventricle. Am J Physiol Heart Circ Physiol. 2001;280:H610–620. doi: 10.1152/ajpheart.2001.280.2.H610

13. Zwanenburg JJM, Götte MJW, Kuijer JPA, Heethaar RM, Rossum ACv, Marcus JT. Timing of cardiac contraction in humans mapped by high-temporal-resolution MRI tagging: early onset and late peak of shortening in lateral wall. American Journal of Physiology-Heart and Circulatory Physiology. 2004;286:H1872–H1880. doi: 10.1152/ajpheart.01047.2003

14. Herman MV, Gorlin R. Implications of left ventricular asynergy. The American Journal of Cardiology. 1969;23:538–547. doi: 10.1016/0002-9149(69)90007-1

15. Herman MV, Heinle RA, Klein MD, Gorlin R, Hammond E, Tsao-Wu N, Levy R. Localized Disorders in Myocardial Contraction. New England Journal of Medicine. 1967;277:222–232. doi: doi:10.1056/NEJM196708032770502

16. Prinzen FW, Augustijn CH, Allessie MA, Arts T, Delhaas T, Reneman RS. The time sequence of electrical and mechanical activation during spontaneous beating and ectopic stimulation. European Heart Journal. 1992;13:535–543.

17. Lyseggen E, Vartdal T, Remme EW, Helle-Valle T, Pettersen E, Opdahl A, Edvardsen T, Smiseth OA. A novel echocardiographic marker of end systole in the ischemic left ventricle: “tug of war” sign. American Journal of Physiology-Heart and Circulatory Physiology. 2009;296:H645–H654. doi: 10.1152/ajpheart.00313.2008

18. Little WC, Reeves RC, Arciniegas J, Katholi RE, Rogers EW. Mechanism of abnormal interventricular septal motion during delayed left ventricular activation. Circulation. 1982;65:1486–1491. doi: doi:10.1161/01.CIR.65.7.1486

19. Russell K, Smiseth OA, Gjesdal O, Qvigstad E, Norseng PA, Sjaastad I, Opdahl A, Skulstad H, Edvardsen T, Remme EW. Mechanism of prolonged electromechanical delay in late activated myocardium during left bundle branch block. American Journal of Physiology-Heart and Circulatory Physiology. 2011;301:H2334–H2343. doi: 10.1152/ajpheart.00644.2011

20. Aalen JM, Remme EW, Larsen CK, Andersen OS, Krogh M, Duchenne J, Hopp E, Ross S, Beela AS, Kongsgaard E, et al. Mechanism of Abnormal Septal Motion in Left Bundle Branch Block: Role of Left Ventricular Wall Interactions and Myocardial Scar. JACC: Cardiovascular Imaging. 2019;12:2402–2413. doi: 10.1016/j.jcmg.2018.11.030

21. Cleland JGF, Daubert JC, Erdmann E, Freemantle N, Gras D, Kappenberger L, Tavazzi L. The effect of cardiac resynchronization on morbidity and mortality in heart failure. New England Journal of Medicine. 2005;352:1539–1549. doi: 10.1056/NEJMoa050496

22. Chitiboi T, Axel L. Magnetic resonance imaging of myocardial strain: A review of current approaches. Journal of Magnetic Resonance Imaging. 2017;46:1263–1280. doi: 10.1002/jmri.25718

23. Nayak KS, Nielsen J-F, Bernstein MA, Markl M, D. Gatehouse P, M. Botnar R, Saloner D, Lorenz C, Wen H, S. Hu B, et al. Cardiovascular magnetic resonance phase contrast imaging. Journal of Cardiovascular Magnetic Resonance. 2015;17:71. doi: 10.1186/s12968-015-0172-7

24. Espe EKS, Aronsen JM, Eriksen M, Sejersted OM, Zhang L, Sjaastad I. Regional Dysfunction After Myocardial Infarction in Rats. Circulation: Cardiovascular Imaging. 2017;10:e005997. doi: doi:10.1161/CIRCIMAGING.116.005997

25. Broch K, Anstensrud AK, Woxholt S, Sharma K, Tøllefsen IM, Bendz B, Aakhus S, Ueland T, Amundsen BH, Damås JK, et al. Randomized Trial of Interleukin-6 Receptor Inhibition in Patients With Acute ST-Segment Elevation Myocardial Infarction. Journal of the American College of Cardiology. 2021;77:1845–1855. doi: doi:10.1016/j.jacc.2021.02.049

26. Blom KB, Bergo KK, Espe EKS, Rosseland V, Grøtta OJ, Mjøen G, Åsberg A, Bergan S, Sanner H, Bergersen TK, et al. Cardiovascular rEmodelling in living kidNey donorS with reduced glomerular filtration rate: rationale and design of the CENS study. Blood Press. 2020;29:123–134. doi: 10.1080/08037051.2019.1684817

27. Reese TG, Feinberg DA, Dou J, Wedeen VJ. Phase contrast MRI of myocardial 3D strain by encoding contiguous slices in a single shot. Magn Reson Med. 2002;47:665–676. doi: 10.1002/mrm.10111

28. Espe EKS, Skårdal K, Aronsen JM, Zhang L, Sjaastad I. A semiautomatic method for rapid segmentation of velocity-encoded myocardial magnetic resonance imaging data. Magn Reson Med. 2017;78:1199–1207. doi: 10.1002/mrm.26486

29. Espe EK, Aronsen JM, Skrbic B, Skulberg VM, Schneider JE, Sejersted OM, Zhang L, Sjaastad I. Improved MR phase-contrast velocimetry using a novel nine-point balanced motion-encoding scheme with increased robustness to eddy current effects. Magn Reson Med. 2013;69:48–61. doi: 10.1002/mrm.24226

30. Johnson KM, Markl M. Improved SNR in phase contrast velocimetry with five-point balanced flow encoding. Magn Reson Med. 2010;63:349–355. doi: 10.1002/mrm.22202

31. Nilsson A, Markenroth Bloch K, Carlsson M, Heiberg E, Ståhlberg F. Variable velocity encoding in a three-dimensional, three-directional phase contrast sequence: Evaluation in phantom and volunteers. J Magn Reson Imaging. 2012;36:1450–1459. doi: 10.1002/jmri.23778

32. Espe EK, Aronsen JM, Eriksen GS, Zhang L, Smiseth OA, Edvardsen T, Sjaastad I, Eriksen M. Assessment of regional myocardial work in rats. Circ Cardiovasc Imaging. 2015;8:e002695. doi: 10.1161/circimaging.114.002695

33. Russell K, Eriksen M, Aaberge L, Wilhelmsen N, Skulstad H, Gjesdal O, Edvardsen T, Smiseth OA. Assessment of wasted myocardial work: a novel method to quantify energy loss due to uncoordinated left ventricular contractions. Am J Physiol Heart Circ Physiol. 2013;305:H996–1003. doi: 10.1152/ajpheart.00191.2013

34. Rice JJ, Wang F, Bers DM, de Tombe PP. Approximate model of cooperative activation and crossbridge cycling in cardiac muscle using ordinary differential equations. Biophys J. 2008;95:2368–2390. doi: 10.1529/biophysj.107.119487

35. Cerqueira MD, Weissman NJ, Dilsizian V, Jacobs AK, Kaul S, Laskey WK, Pennell DJ, Rumberger JA, Ryan T, Verani MS. Standardized myocardial segmentation and nomenclature for tomographic imaging of the heart. A statement for healthcare professionals from the Cardiac Imaging Committee of the Council on Clinical Cardiology of the American Heart Association. Circulation. 2002;105:539–542. doi: 10.1161/hc0402.102975

36. Cazorla O, Lacampagne A. Regional variation in myofilament length-dependent activation. Pflugers Arch. 2011;462:15–28. doi: 10.1007/s00424-011-0933-6

37. Piacentino V, Weber CR, Chen X, Weisser-Thomas J, Margulies KB, Bers DM, Houser SR. Cellular Basis of Abnormal Calcium Transients of Failing Human Ventricular Myocytes. Circulation Research. 2003;92:651–658. doi: doi:10.1161/01.RES.0000062469.83985.9B

38. Myles RC, Bernus O, Burton FL, Cobbe SM, Smith GL. Effect of activation sequence on transmural patterns of repolarization and action potential duration in rabbit ventricular myocardium. American Journal of Physiology-Heart and Circulatory Physiology. 2010;299:H1812–H1822. doi: 10.1152/ajpheart.00518.2010

39. Wasserstrom JA, Shiferaw Y, Chen W, Ramakrishna S, Patel H, Kelly JE, O’Toole MJ, Pappas A, Chirayil N, Bassi N, et al. Variability in timing of spontaneous calcium release in the intact rat heart is determined by the time course of sarcoplasmic reticulum calcium load. Circ Res. 2010;107:1117–1126. doi: 10.1161/circresaha.110.229294

40. Davis JS, Hassanzadeh S, Winitsky S, Lin H, Satorius C, Vemuri R, Aletras AH, Wen H, Epstein ND. The Overall Pattern of Cardiac Contraction Depends on a Spatial Gradient of Myosin Regulatory Light Chain Phosphorylation. Cell. 2001;107:631–641. doi: 10.1016/S0092-8674(01)00586-4

41. Louch WE, Bito V, Heinzel FR, Macianskiene R, Vanhaecke J, Flameng W, Mubagwa K, Sipido KR. Reduced synchrony of Ca2+ release with loss of T-tubules-a comparison to Ca2+ release in human failing cardiomyocytes. Cardiovasc Res. 2004;62:63–73. doi: 10.1016/j.cardiores.2003.12.031

42. Frisk M, Le C, Shen X, Røe ÅT, Hou Y, Manfra O, Silva GJJ, Hout Iv, Norden ES, Aronsen JM, et al. Etiology-Dependent Impairment of Diastolic Cardiomyocyte Calcium Homeostasis in Heart Failure With Preserved Ejection Fraction. Journal of the American College of Cardiology. 2021;77:405–419. doi: doi:10.1016/j.jacc.2020.11.044

43. Katz AM, Katz PB. Homogeneity out of heterogeneity. Circulation. 1989;79:712–717. doi: 10.1161/01.cir.79.3.712

44. Lang RM, Badano LP, Mor-Avi V, Afilalo J, Armstrong A, Ernande L, Flachskampf FA, Foster E, Goldstein SA, Kuznetsova T, et al. Recommendations for cardiac chamber quantification by echocardiography in adults: An update from the American society of echocardiography and the European association of cardiovascular imaging. European Heart Journal Cardiovascular Imaging. 2015;16:233–271. doi: 10.1093/ehjci/jev014

45. Smedsrud MK, Sarvari S, Haugaa KH, Gjesdal O, Ørn S, Aaberge L, Smiseth OA, Edvardsen T. Duration of Myocardial Early Systolic Lengthening Predicts the Presence of Significant Coronary Artery Disease. Journal of the American College of Cardiology. 2012;60:1086–1093. doi: 10.1016/j.jacc.2012.06.022

46. Stankovic I, Prinz C, Ciarka A, Daraban AM, Kotrc M, Aarones M, Szulik M, Winter S, Belmans A, Neskovic AN, et al. Relationship of visually assessed apical rocking and septal flash to response and long-term survival following cardiac resynchronization therapy (PREDICT-CRT). European Heart Journal – Cardiovascular Imaging. 2015;17:262–269. doi: 10.1093/ehjci/jev288

47. Brainin P. Myocardial Postsystolic Shortening and Early Systolic Lengthening: Current Status and Future Directions. Diagnostics (Basel). 2021;11. doi: 10.3390/diagnostics11081428

48. Brainin P, Biering-Sørensen SR, Møgelvang R, Jensen JS, Biering-Sørensen T. Duration of early systolic lengthening: prognostic potential in the general population. Eur Heart J Cardiovasc Imaging. 2020;21:1283–1290. doi: 10.1093/ehjci/jez262

49. Brainin P, Haahr-Pedersen S, Olsen FJ, Holm AE, Fritz-Hansen T, Jespersen T, Gislason G, Biering-Sørensen T. Early Systolic Lengthening in Patients With ST&#x2010;Segment&#x2013;Elevation Myocardial Infarction: A Novel Predictor of Cardiovascular Events. Journal of the American Heart Association. 2020;9:e013835. doi: doi:10.1161/JAHA.119.013835

50. Skulstad H, Edvardsen T, Urheim S, Rabben SI, Stugaard M, Lyseggen E, Ihlen H, Smiseth OA. Postsystolic Shortening in Ischemic Myocardium. Circulation. 2002;106:718–724. doi: doi:10.1161/01.CIR.0000024102.55150.B6

51. Takayama M, Norris RM, Brown MA, Armiger LC, Rivers JT, White HD. Postsystolic shortening of acutely ischemic canine myocardium predicts early and late recovery of function after coronary artery reperfusion. Circulation. 1988;78:994–1007. doi: 10.1161/01.cir.78.4.994

52. Lyseggen E, Skulstad H, Helle-Valle T, Vartdal T, Urheim S, Rabben SI, Opdahl A, Ihlen H, Smiseth OA. Myocardial strain analysis in acute coronary occlusion: a tool to assess myocardial viability and reperfusion. Circulation. 2005;112:3901–3910. doi: 10.1161/circulationaha.105.533372

53. Glass L. Synchronization and rhythmic processes in physiology. Nature. 2001;410:277–284. doi: 10.1038/35065745

54. Goldberger AL, Amaral LAN, Hausdorff JM, Ivanov PC, Peng C-K, Stanley HE. Fractal dynamics in physiology: Alterations with disease and aging. Proceedings of the National Academy of Sciences. 2002;99:2466–2472. doi: doi:10.1073/pnas.012579499

55. Braunwald E, Ross J, Sonnenblick EH. Mechanisms of Contraction of the Normal and Failing Heart. New England Journal of Medicine. 1967;277:794–800. doi: doi:10.1056/NEJM196710122771505

56. Kentish JC, ter Keurs HE, Ricciardi L, Bucx JJ, Noble MI. Comparison between the sarcomere length-force relations of intact and skinned trabeculae from rat right ventricle. Influence of calcium concentrations on these relations. Circulation Research. 1986;58:755–768. doi: doi:10.1161/01.RES.58.6.755

57. Bers DM. Cardiac excitation–contraction coupling. Nature. 2002;415:198–205. doi: 10.1038/415198a

58. Najafi A, Sequeira V, Kuster DW, van der Velden J. β-adrenergic receptor signalling and its functional consequences in the diseased heart. Eur J Clin Invest. 2016;46:362–374. doi: 10.1111/eci.12598

59. Song LS, Wang SQ, Xiao RP, Spurgeon H, Lakatta EG, Cheng H. beta-Adrenergic stimulation synchronizes intracellular Ca(2+) release during excitation-contraction coupling in cardiac myocytes. Circ Res. 2001;88:794–801. doi: 10.1161/hh0801.090461

60. Amoni M, Vermoortele D, Ekhteraei-Tousi S, Doñate Puertas R, Gilbert G, Youness M, Thienpont B, Willems R, Roderick HL, Claus P, et al. Heterogeneity of Repolarization and Cell-Cell Variability of Cardiomyocyte Remodeling Within the Myocardial Infarction Border Zone Contribute to Arrhythmia Susceptibility. Circ Arrhythm Electrophysiol. 2023;16:e011677. doi: 10.1161/circep.122.011677

61. Weber KT, Sun Y, Bhattacharya SK, Ahokas RA, Gerling IC. Myofibroblast-mediated mechanisms of pathological remodelling of the heart. Nature Reviews Cardiology. 2013;10:15–26. doi: 10.1038/nrcardio.2012.158

62. Lou Q, Fedorov VV, Glukhov AV, Moazami N, Fast VG, Efimov IR. Transmural heterogeneity and remodeling of ventricular excitation-contraction coupling in human heart failure. Circulation. 2011;123:1881–1890.

63. Neubauer S. The Failing Heart — An Engine Out of Fuel. New England Journal of Medicine. 2007;356:1140–1151. doi: doi:10.1056/NEJMra063052

64. Haertter D, Hauke L, Driehorst T, Nishi K, Zimmermann W-H, Schmidt CF. Stochastic tug-of-war among sarcomeres mediates cardiomyocyte response to environmental stiffness. In: eLife Sciences Publications, Ltd; 2024.

65. Kobirumaki-Shimozawa F, Oyama K, Nakanishi T, Ishiwata S, Fukuda N. Asynchronous movement of sarcomeres in myocardium under living conditions: role of titin. Front Physiol. 2024;15:1426545. doi: 10.3389/fphys.2024.1426545

66. Li J, Sundnes J, Hou Y, Laasmaa M, Ruud M, Unger A, Kolstad TR, Frisk M, Norseng PA, Yang L, et al. Stretch Harmonizes Sarcomere Strain Across the Cardiomyocyte. Circulation Research. 2023;133:255–270. doi: doi:10.1161/CIRCRESAHA.123.322588

67. Ariga R, Tunnicliffe EM, Manohar SG, Mahmod M, Raman B, Piechnik SK, Francis JM, Robson MD, Neubauer S, Watkins H. Identification of Myocardial Disarray in Patients With Hypertrophic Cardiomyopathy and Ventricular Arrhythmias. J Am Coll Cardiol. 2019;73:2493–2502. doi: 10.1016/j.jacc.2019.02.065

68. Camici PG, Crea F. Coronary Microvascular Dysfunction. New England Journal of Medicine. 2007;356:830–840. doi: doi:10.1056/NEJMra061889

69. Kuppe C, Ramirez Flores RO, Li Z, Hayat S, Levinson RT, Liao X, Hannani MT, Tanevski J, Wünnemann F, Nagai JS, et al. Spatial multi-omic map of human myocardial infarction. Nature. 2022;608:766–777. doi: 10.1038/s41586-022-05060-x

